# Interactive Physical Activity Apps: Do the ABACUS and the MARS Measure Up? A Descriptive Analysis of Behaviour Change Taxonomies

**DOI:** 10.64898/2026.02.18.26346599

**Authors:** Elaine M. Ori, Courtney Baay, Manuel Ester, Ann M Toohey

## Abstract

The ubiquitous use of digital tools may be beneficial for improving physical activity across diverse populations. It remains unknown however, how publicly available, cost-free physical activity apps adhere to behaviour change techniques, and how users rate these apps. To explore the number of publicly available physical activity apps and relationships among behaviour science techniques, subjective quality, and user ratings. Exploratory content analysis of 17 apps meeting inclusion criteria. The App Behaviour Change Scale (ABACUS) and Mobile App Rating Scale (MARS) were used to code each downloaded app for behaviour change techniques, app functionality, and subjective quality. App store user ratings were also collected along with descriptive data about each app. All apps were commercially affiliated, targeted adult populations, and centered on changing behaviour, setting goals, and addressing physical health. No apps addressed all 21 ABACUS items; apps included 12.8 ± 2.4 indicators, ranging from 8-18 indicators. The three most common ABACUS indicators were: i) collection of baseline information, ii) instructional PA content, and iii) ability for app to give user feedback. The three least common ABACUS indicators were: i) ability to export data, ii) consequences for physical activity dis/continuance, and iii) allows for planning of barriers. No apps included all 12 MARS focus areas; 94.1% of apps allowed goal setting, 58.8% addressed physical health, and 41.2% included a mindfulness focus. Linear regressions explored relationships for app user ratings; aggregated MARS domains accounted for 54% of the variance. Publicly available physical activity apps may be a useful approach to improving physical activity uptake and adherence among harder-to-reach populations including low socioeconomic status groups. App developers should consider incorporating more behaviour change techniques within cost-free apps to improve user uptake and ultimately improve physical activity associated health outcomes.

**Author Summary:** Digital technology proliferates all facets of life and populations, and may contribute to improved health behaviours including physical activity. However, access to supportive technology may be limited by cost for example, as many popular physical activity apps require paid subscriptions. It is unknown whether cost-free physical activity apps adhere to behaviour change recommendations and how these apps are rated by users. This research explored cost-free, publicly available physical activity apps and their respective relationships with behaviour change techniques as well as app-store user ratings. Only 17 apps met inclusion criteria, and were compared against one behaviour change scale and one app quality scale. All apps had commercial motivations and focused on physical activity for adult populations. Most commonly, apps collected user info at baseline, provided physical activity instructional content, and provided feedback to users. Apps were generally rated positively by users based on app-store star ratings. Cost-free physical activity apps may be useful tools for users looking to improve physical activity for individuals who are limited by their socioeconomic situation. However, greater emphasis on evidence-based behaviour change approaches may be necessary to improve health outcomes for users.

---

The benefits of physical activity are widely known and include reduction in all-cause mortality, reduced risk for cancers, type II diabetes, arthritis, and improved cognitive function(1–3). Despite physical activity (PA) guidelines recommending that adults accumulate at least 150 minutes of moderate-vigorous aerobic PA plus engage in two resistance training bouts per week(4), evidence suggests that any amount of PA improves health and lowers disease risk(1,3). Yet waning PA levels are a leading contributor to all-cause mortality and disease, as well as significant health system and economic burden(5–8). Defined as any energy-expending movement, the term ‘physical activity’ is often conflated with ‘fitness’ and ‘exercise’(9). While exercise is a structured form of PA that is typically done for the purpose of improving skill or physical fitness, fitness refers to PA intended to improve vigor, which may include cardiorespiratory endurance, muscular strength or endurance, body composition, and flexibility (9). Despite the colloquial use of these terms interchangeably, emphasis has historically been placed on engagement in traditional exercise and fitness activities to maintain an ideal physique and for athletic pursuits(10–13). Consequently, use of digital PA tools is often focused on providing guidance, instruction, and education about various exercise modalities including yoga, running, flexibility, and strength training, for improving or tracking fitness.

Digital exercise and fitness tools have shown promise as effective interventions for improving PA among a variety of populations(14–16). The proliferation of user-supported digital advice available includes websites, downloadable applications (or “apps”), and wearables(17). Use of fitness apps has been identified as the second most prevalent fitness trend in 2025(18). Among digital health apps, PA-based apps (i.e., exercise, fitness) remain one of the largest sub-sets, comprising up to 30% of the market(19,20). Approximately 23.33 million fitness apps were downloaded onto personal electronic devices in 2024, generating $3.88 billion (USD) in revenue for the same year(21). However, gaps exist, as behaviour change may require tailoring content and outcomes based on individual health status and personal goals. Digital behaviour change interventions may be more effective for healthy rather than unhealthy populations(14). Behavior change research suggests that PA-based apps should include at least one element of behaviour change taxonomy(20). However, most apps also have commercial affiliations and may not be developed using behaviour science approaches(20). The use of wearable technology (i.e., watches, tracking monitors), smartphone sensors, and devices is often included in PA apps to track app users’ PA (16). Qualitative studies have identified app-assisted behaviour tracking as a desirable feature of using health apps(22) however, high costs associated with tracking devices such as smart watches may present barriers for low-socioeconomic groups. Nonetheless, individualized behaviour change support, when using digital apps, has been effective at improving health behaviour change(16), and identified as desirable features for PA app users (22) .

Knowledge of successful behaviour change techniques, coupled with users’ experiences of health-based app use, has contributed to the development of behaviour change taxonomies(23,24). When validated against various fitness apps, it appears that incorporating some or all behaviour change taxonomies may improve app efficacy for behaviour change(25). Yet many apps lack evidence-informed behaviour change techniques(26). Additionally, despite the high-volume of exercise and fitness apps available for mobile download, no known studies have examined the relationship between app-based user ratings and behavioural taxonomies for PA change. The objective of the study therefore, was threefold: first, to analyze the number of active, cost-free digital applications available to the public and to assess the extent to which each application incorporates behaviour change techniques; second, to explore the relationships between app user ratings and behavior change techniques; and third, to assess subjective quality of the apps for behaviour change principles. Given the exploratory nature of our research aims, no *a priori* hypotheses are stated.

## Methods

This digital environmental scan systematically identified and characterized apps that promote PA among adults. To address the research objectives, the digital environmental scan focused on popular, cost-free PA-promoting apps available in the Google Play (Android) and Apple iTunes stores. The App Behaviour Change Scale (ABACUS)(24) was used, as one taxonomy that has been shown effective for classifying the behaviour change potential of PA apps. Incorporating measures identified as integral for initiating behaviour change, the ABACUS is a 21-item scale that classifies the behaviour change potential of apps. The ABACUS includes measures of knowledge and information, goals and planning, feedback and monitoring, and actions related to the target behaviour. The Mobile App Rating Scale (MARS)(27) has also been suggested as an important tool for assessing mobile app quality. The MARS is an additional app evaluation tool, developed to classify app quality based on 23 items across four objective scales that include app engagement, functionality, aesthetics, and quality of information. The MARS includes an additional subjective scale, assessing the subjective quality of an app.

### Search Procedure

Our comprehensive search used an approach to data collection, guided by previous research examining digital health interventions(28–30). Thus, apps were explored using three key terms, “physical activity”, “fitness”, and “exercise”, as was done in prior work(28,29). We searched Google Play and Apple iTunes stores, to capture recent, current returns. Two reviewers independently conducted parallel searches using the main search bar within the Google Play and Apple iTunes stores, with one reviewer specifically assigned to Google Play and the other to Apple iTunes. A sample of up to 100 apps per app store for each search term was collected(30,31). Apps requiring paid subscriptions to access interactive components were excluded. Searches were initiated and completed on two consecutive days by two independent reviewers. This approach mitigated variance caused by temporal spacing, as app popularity can fluctuate daily based on usage(32–34).

### Screening Procedure

The names and descriptions, as published by the app stores, of each identified app was catalogued. Two reviewers independently assessed each app against the *a priori* inclusion and exclusion criteria based on its title and description. After calculating inter-rater reliability, the primary reviewer completed title and description assessment of the remaining apps, resulting in a final sample of apps. The final sample of apps was then downloaded by the two reviewers onto either an Apple iPhone or Samsung Galaxy Android phone. If an app was available on both Google Play and Apple iTunes, it was defaulted to the reviewer with Google Play Store access for download and assessment. Each app was left running in the background for two days to capture any pop-ups, notifications, and/or reminders. Reviewers collected screenshots and assessed each app based on a pre-defined *pro forma* extraction table. After two days, the app was removed.

### Data Extraction and Coding

Data extraction and coding were conducted using two validated scales: the App Behaviour Change Scale (ABACUS)(24) to assess each app’s potential for behavior change features, and the Mobile App Rating Scale (MARS)(27) to evaluate each app’s quality, as described above. Two researchers systematically coded a 33% subsample of the included apps using both the ABACUS and MARS scales. Given the small inter-rater sample size (*n* = 6) percentage agreement was calculated as 83.3% - 100.0% agreement suggesting near-perfect to perfect agreement(36) for dichotomous MARS and ABACUS items. Paired samples t-tests were used to determine agreement for MARS scale items, all non-significant indicating agreement between raters. All discrepancies were resolved through discussion between the two researchers. Subsequently, one independent researcher extracted and coded the remaining apps.

## Data Analysis

Descriptive statistics were calculated to summarize the data, including means and standard deviations for normally distributed continuous variables, medians and interquartile ranges for non-normal continuous variables, and percentages and frequencies for categorical variables. Pearson correlation coefficients were calculated to explore potential relationships between the app ratings and their respective ABACUS and MARS ratings. Linear regressions were used to further examine relationships between app ratings and ABACUS and MARS ratings. Simple linear regressions included app rating as the dependent variable, with individual domain scores and overall scores for both instruments considered as independent variables. Multiple linear regressions were then used to examine whether app rating was associated with all five MARS domains or all four ABACUS domains, as independent variables. Lastly, the regression analyses were repeated with MARS subjective quality as the dependent variable, to examine whether findings differed when using app store ratings, compared to expert reviewers’ MARS subjective quality ratings. All analyses were completed using R (Version 4.5.0, Mac OS) with an alpha of 0.05.

## Results

An initial sample were collected from each of the Google Play (*n* = 300) and Apple iTunes (*n* = 300) stores. After removing duplicates (n = 242), a final list of apps was generated. Inter-rater reliability for screening was compared for 10% of the final sample (*n* = 358) and calculated Cohen’s Kappa, к = 0.88, P <0.001. One coder completed the remaining sample coding noting additional apps that did not meet inclusion criteria, such as apps did not include an element of customization/personalization or interactivity (*n* = 173), apps were not PA-focused (*n* = 71), or that apps required wearables (*n* = 16). A final sample of apps (*n*=74) were downloaded and again compared against inclusion criteria, with additional apps being excluded (*n* = 55) for reasons such as requiring paid access (*n*= 47), or being inactive (*n* = 5); see Figure 1. A total of 19 apps were included for data extraction; data were ultimately extracted from a total of 17 apps, as two of the original 19 ceased to exist during the course of our study.

**Figure 1.**
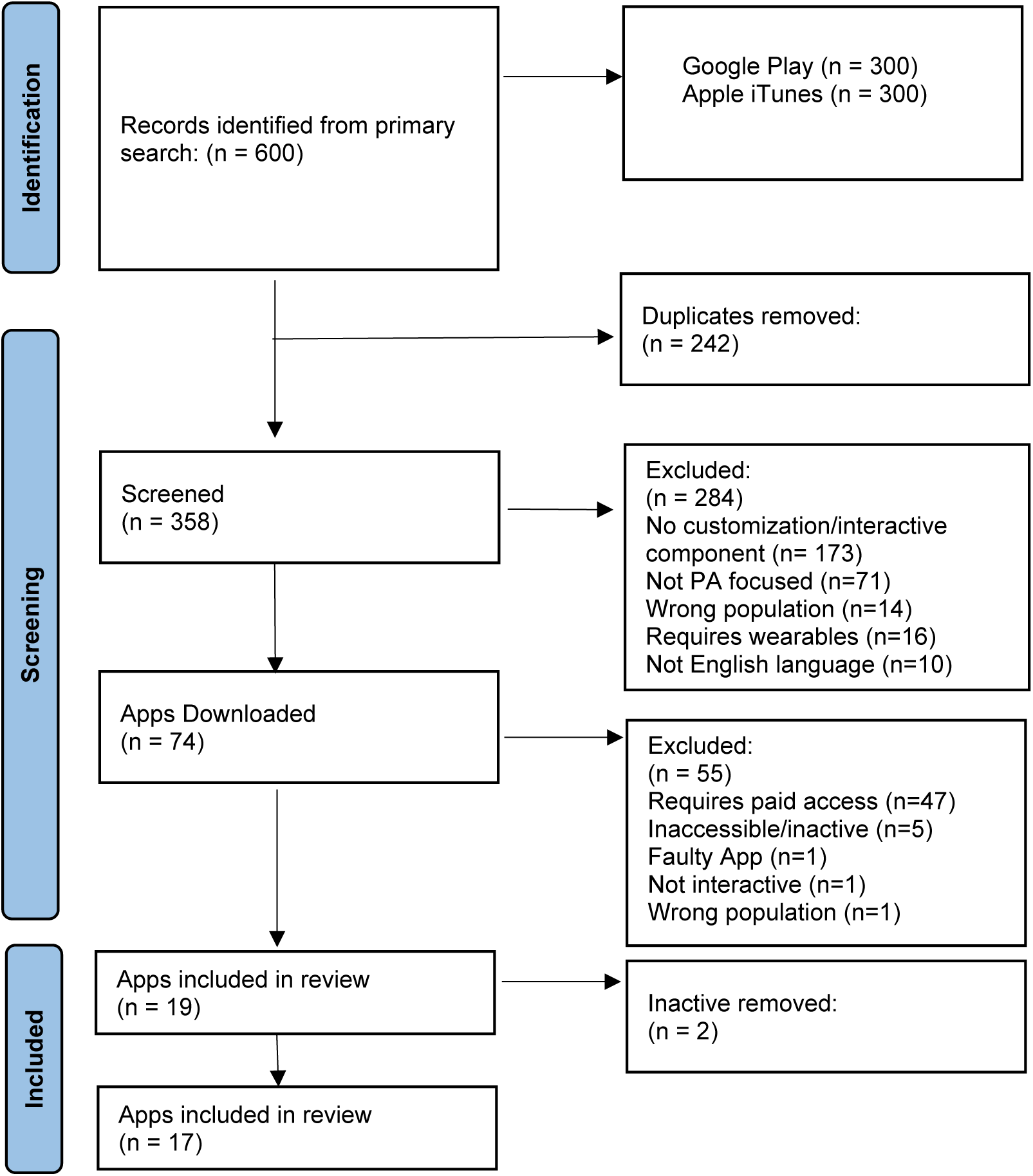
PRISMA Diagram of Apps

Apps were associated with a variety of developers, and 12 apps were at a version of 1.1 or greater. Android-available apps were more prevalent (n = 13, 76.5%) with four apps (23.5%) available only via Apple iPhone. Downloads for the apps ranged from 50,000 to more than 100 million, with numbers unavailable for four apps. Nine (52.9%) apps were updated within one year of review. User ratings were based on a score range of 0-5, with 5 denoting user-rated excellence, µ = 4.5 ± 0.4 (n = 16). Descriptive app characteristics are found in Table 2.

**Table 1.** Eligibility Criteria.

| Inclusion Criteria | Exclusion Criteria |
| --- | --- |
| All content was in English | Apps that targeted activities like glucose monitoring and facial exercise |
| PA* was the primary target behavior<br>[*Any bodily movement produced by skeletal muscles that results in energy expenditure beyond sedentary behaviour, encompassing activities such as rehabilitative exercise, resistance training, cardiovascular training, sport-specific training, etc.(35)] | Apps that offered one-way direct-to-consumer information or workout repositories |
| The target population was 18 years and over. | Apps that exclusively tracked PA, such as step counts or activity logs. |
| The interface was two-way and interactive, allowing users to provide input and receive output (e.g., progress reports, custom workout plans, community platforms). | Apps that *required* the use of wearables (e.g., smartwatch) or any kind of external equipment (e.g., dumbbells) |

**Table 2.** Descriptive Characteristics for Physical Activity Apps.

| **App Name** | **Developer** | **Version** | **Rating this version\*** | **Number of Downloads** | **Number of ratings this version** | **Platform (iPhone, Android)** |
| --- | --- | --- | --- | --- | --- | --- |
| Decathlon Coach | Decathlon | 2.29.0 | 4 | 5million+ | 80000 | Android |
| Fitness Online | Fitness Online LLC | 2.18.1 | 4.7 | 5million+ | 54000 | Android |
| Nike Train Club | Nike, Inc. | 6.56.0 | 4.4 | 10million+ | 360000 | Android |
| Bodbot | BodBot | 6.187 | 4.6 | 5million+ | 53000 | Android |
| Anytime Fitness | Anytime Fitness, LLC, | 2.47.3 | 3.3 | 500000+ | 1000 | Android |
| Fitness Coach | Leap Fitness Group | 1.1.9 | 4.8 | 10million+ | 255000 | Android |
| Live Well | Zurich LiveWell Services & Solutions | unavailable | unavailable | unavailable | 0 | iPhone |
| Home Workout: Fitness No Tools | LifeBuddy | unavailable | 4.6 | 1million+ | 12000 | Android |
| Home Workout - No Equipments | Leap Fitness Group | unavailable | 4.9 | 100million+ | 3000000 | Android |
| Fittr: Fitness & Weight Loss | Fitter - Fitness 7 Personal Training | 8.7.5 | 4.6 | 1million+ | 20000 | Android |
| SHRED: Gym & Home Workout | Shred Labs,LLC | v2.5.7 | 4.2 | 50000+ | 978 | Android |
| Booty Coach - Workout+Fitness | Fast Builder Limited | 6.1.6 | 4.8 | unavailable | 4900 | iPhone |
| Fitness App: Home Gym Workout | ITPlus LLC | unavailable | 4.7 | unavailable | 3400 | iPhone |
| Life Time Digital | Life Time, Inc | 6.0.0 | 4.8 | 500000 | 11000 | Android |
| Fresh Tri: Habits & Mindset | Fresh Tri | 2.23.0 | 4.4 | 500000+ | 2000 | Android |
| Madbarz: Bodyweight Workout | MadBarz Group | 6.8.5 | 4.6 | unavailable | 177 | iPhone |
| FitOn Workouts & Fitness Plans | FitOn | 6.6.0 | 4.5 | editor's choice' no number reported | 92000 | Android |

\*Ratings based on five stars maximum where 5 = excellent

### ABACUS Results

A detailed overview of ABACUS ratings is shown in Table 3. In total, scores ranged from 8-18/21 (38.1-85.7%), with an average of 12.8±2.4 (60.8±11.6%). None of the apps addressed all 21 ABACUS indicators, with apps ranging from 8 indicators addressed (Nike Training Club app) to 18 indicators addressed (Fresh Tri: Habits & Mindset app). Average scores for individual ABACUS categories ranged from 57.8±17.8% (Category 4: Actions) to 64.7±24.9% (Category 2: Goals & Planning). Seven individual ABACUS items were present in all apps: 1.3 Collection of baseline information, 1.4 Instructional PA content, 3.4 Ability for app to give user feedback, 3.7 Provides encouragement, 4.1 Has reminders, 4.2 Encourages positive habit formation, 4.3 Allows for practice + daily activities. Meanwhile, the least common features were 3.5 Ability to Export Data (0/17, 0%), 1.5 Consequences for PA dis/continuance (1/17, 5.9%), and 4.4 Allows for Planning of Barriers (2/17, 11.8%).

**Table 3.** Matrix of ABACUS Results.

| App | 1. Knowledge & Information |  |  |  |  | 2. Goals & Planning |  |  | 3. Feedback & Monitoring |  |  |  |  |  |  | 4. Actions |  |  |  |  |  |
| --- | --- | --- | --- | --- | --- | --- | --- | --- | --- | --- | --- | --- | --- | --- | --- | --- | --- | --- | --- | --- | --- |
|  | 1.1 Personalization and Customizable Features | 1.2 Consistent with national PA guidelines/evidence of expertise | 1.3 Collection of baseline information | 1.4 Instructional PA content | 1.5 Consequences for PA dis/continuance | 2.1 Asks willingness for behavior change | 2.2 Allows for goal setting | 2.3 Ability to review, update, change goals. | 3.1 Gives user ability to understand current status vs future goals | 3.2 Ability for user to self-monitor behaviour | 3.3 Ability to share behaviour with others | 3.4 Ability for app to give user feedback | 3.5 Ability to export app data | 3.6 Provides material incentive | 3.7 Provides encouragement | 4.1 Has reminders | 4.2 Encourages positive habit formation | 4.3 Allows for practice + daily activities | 4.4 Allows for planning of barriers | 4.5 Makes suggestions for changing environment | 4.6 Assists with distraction/avoidance |
| Decathlon Coach | X | X | X | X | X |  | X | X |  | X |  | X |  |  | X | X | X | X |  | X | X |
| Fitness Online | X |  | X | X |  |  | X | X | X | X | X | X |  |  | X | X | X | X |  |  |  |
| Nike Training Club |  |  | X | X |  |  |  |  |  |  | X | X |  |  | X | X | X | X |  |  |  |
| Bodbot | X |  | X | X |  |  | X | X |  | X |  | X |  |  | X | X | X | X |  |  |  |
| Anytime Fitness | X |  | X | X |  |  | X | X | X | X | X | X |  |  | X | X | X | X |  |  |  |
| Fitness Coach | X |  | X | X |  |  | X | X | X | X |  | X |  |  | X | X | X | X |  |  |  |
| Live Well | X |  | X | X |  |  | X | X | X | X |  | X |  | X | X | X | X | X |  |  |  |
| Home Workout: Fitness No Tools | X |  | X | X |  |  | X | X |  | X | X | X |  |  | X | X | X | X |  |  |  |
| Home Workout - No Equipments | X |  | X | X |  | X | X | X | X | X | X | X |  | X | X | X | X | X |  |  |  |
| Fittr: Fitness & Weight Loss | X |  | X | X |  |  | X | X | X | X | X | X |  | X | X | X | X | X |  |  |  |
| SHRED: Gym & Home Workout | X |  | X | X |  |  | X | X | X | X | X | X |  |  | X | X | X | X |  |  |  |
| Booty Coach - Workout+Fitness | X |  | X | X |  | X | X | X | X | X |  | X |  |  | X | X | X | X |  |  |  |
| Fitness App: Home Gym Workout | X |  | X | X |  |  | X |  |  |  | X | X |  |  | X | X | X | X |  |  |  |
| Life Time Digital | X |  | X | X |  |  | X |  |  | X |  | X |  |  | X | X | X | X |  |  |  |
| Fresh Tri: Habits & Mindset | X | X | X | X |  | X | X | X | X | X | X | X |  |  | X | X | X | X | X | X | X |
| Madbarz: Bodyweight Workout | X |  | X | X |  |  | X | X |  |  | X | X |  |  | X | X | X | X |  |  |  |
| FitOn Workouts & Fitness Plans | X |  | X | X |  |  | X | X | X | X | X | X |  |  | X | X | X | X | X | X | X |

Most apps addressed Category 1: Knowledge & Information via collecting baseline information and offering users instructional PA content (both 100%; n = 17). Addressing Goals & Planning appeared in 94.1% (n = 16) of the apps, which included goal setting opportunities. Only 17.6% (n = 3) apps asked users about their willingness to change behaviour. Category 3: Feedback and Monitoring was addressed by all 17 apps by (i) 3.4 Ability for app to Give Feedback, and (ii) 3.7 Provides Encouragement. Two apps included all indicators for Category 4: Actions, and all apps addressed this category by providing users with reminders, encouraging positive habit formation, and allowing practice and daily activities.

### MARS Results

MARS focus areas, theoretical background, affiliations, age group, and technical aspects of included apps, are outlined in Table 4. All apps (17/17, 100%) had commercial affiliations and focused on adults, with one app (1/17, 5.9%) specifically targeted towards young adults between 18-25 years old. Behavior change (17/17, 100%), goal setting (16/17, 94.1%), and physical health (10/17, 58.8%) were the most common focus areas, although 41.2% (n=7) apps also included a mindfulness focus. Seven of the 12 MARS focus areas were represented in at least one app. Whereas feedback, information/education, monitoring/tracking, and advice/tips/skill training were featured as strategies for all 17 apps, none used evidence-based therapeutic techniques such as Cognitive Behavioral Therapy, Acceptance commitment therapy, or strengths-based approaches. More than half of all apps used technical features such as reminders (17/17, 100%), password protection (13/17, 76.5%), and login requirements (10/17, 58.8%).

**Table 4.**
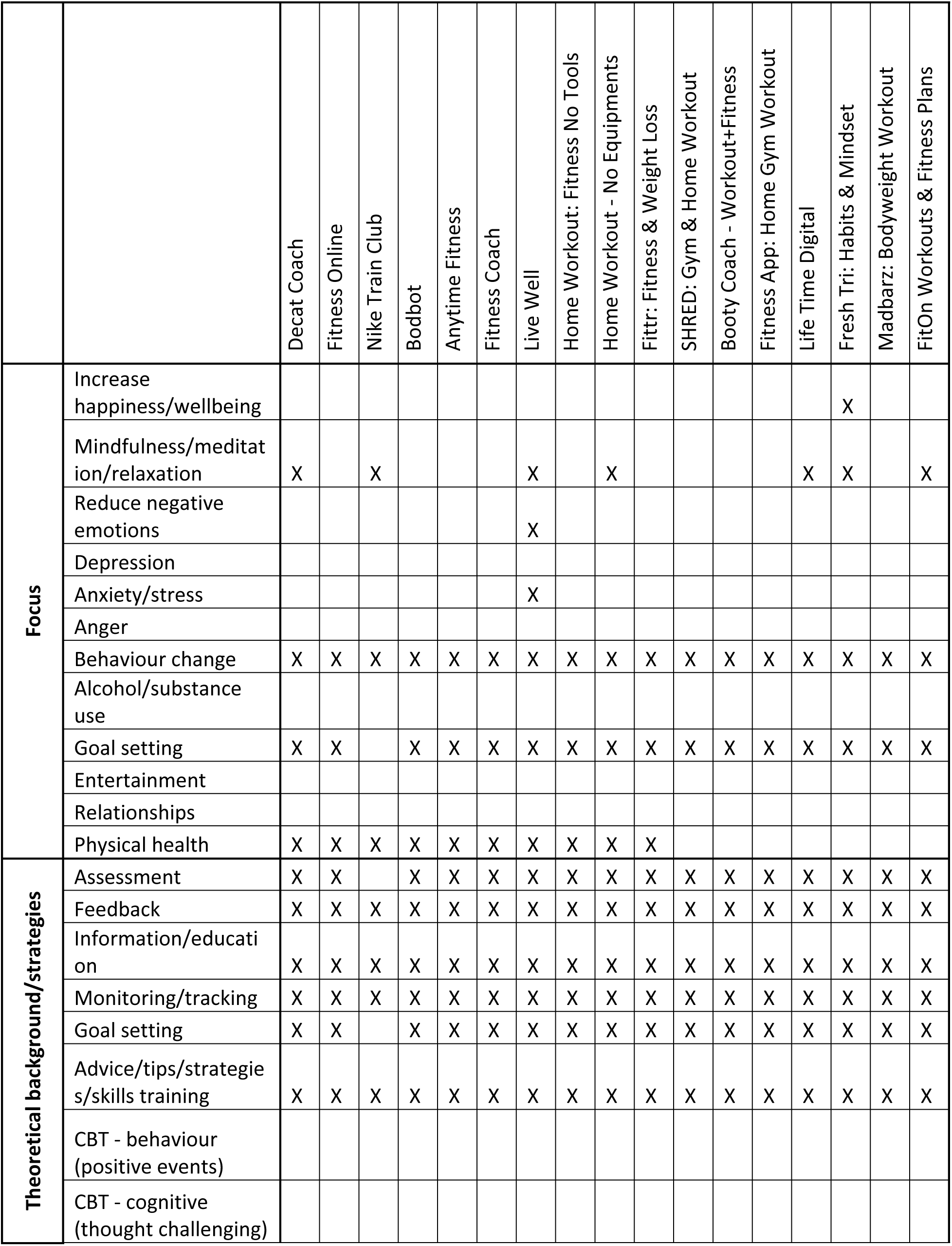
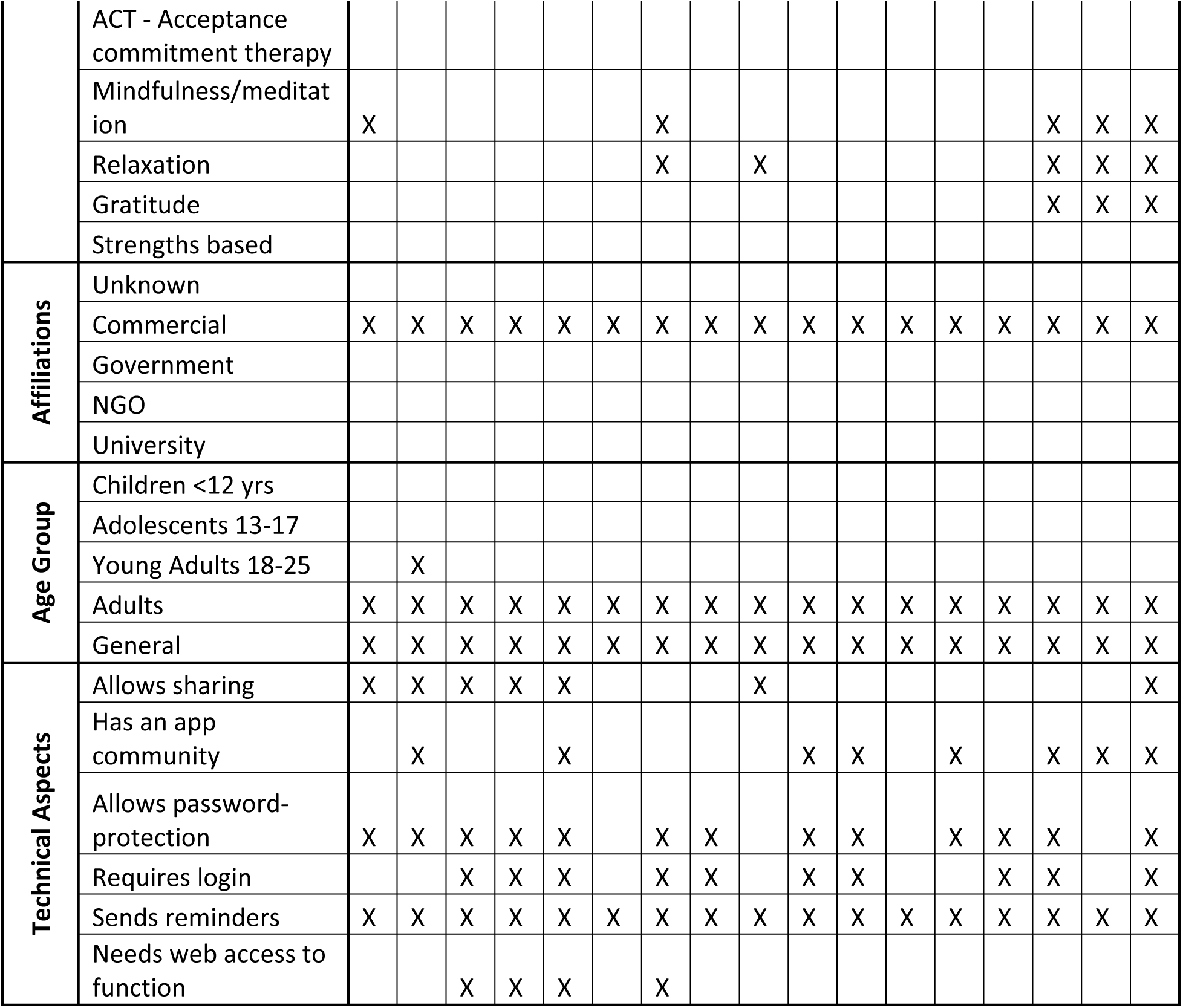
Matrix of MARS results.

All apps were commercially affiliated. All apps were classified as being focused on behaviour change according to the MARS, which also allows for multi-focus classification. All apps included the theoretical strategies of assessing user ability/interest, providing users feedback, providing users information or education, allowing users to track or monitor their activity, allowed users to set goals, and provided advice or skills training. All apps also sent reminders to users while 47.1% (n=8) apps created an app community for users to engage with, and 41.2% (n=7) apps allowed users to share their progress with others. A full matrix of MARS indicators per app is provided in Table 4.

Figure 2 presents histograms of mean MARS ratings for each section, as well as the overall quality score. The lowest mean score was for subjective quality, with a range of 2.0-4.5 (µ = 3.5 ± 0.7). The highest mean score was for functionality, with a range of 3.5-4.8 (µ = 4.2 ± 0.4). The mean overall app quality score, combining engagement, functionality, aesthetics, and information, was µ = 4.0 ± 0.4.

**Figure 2.**
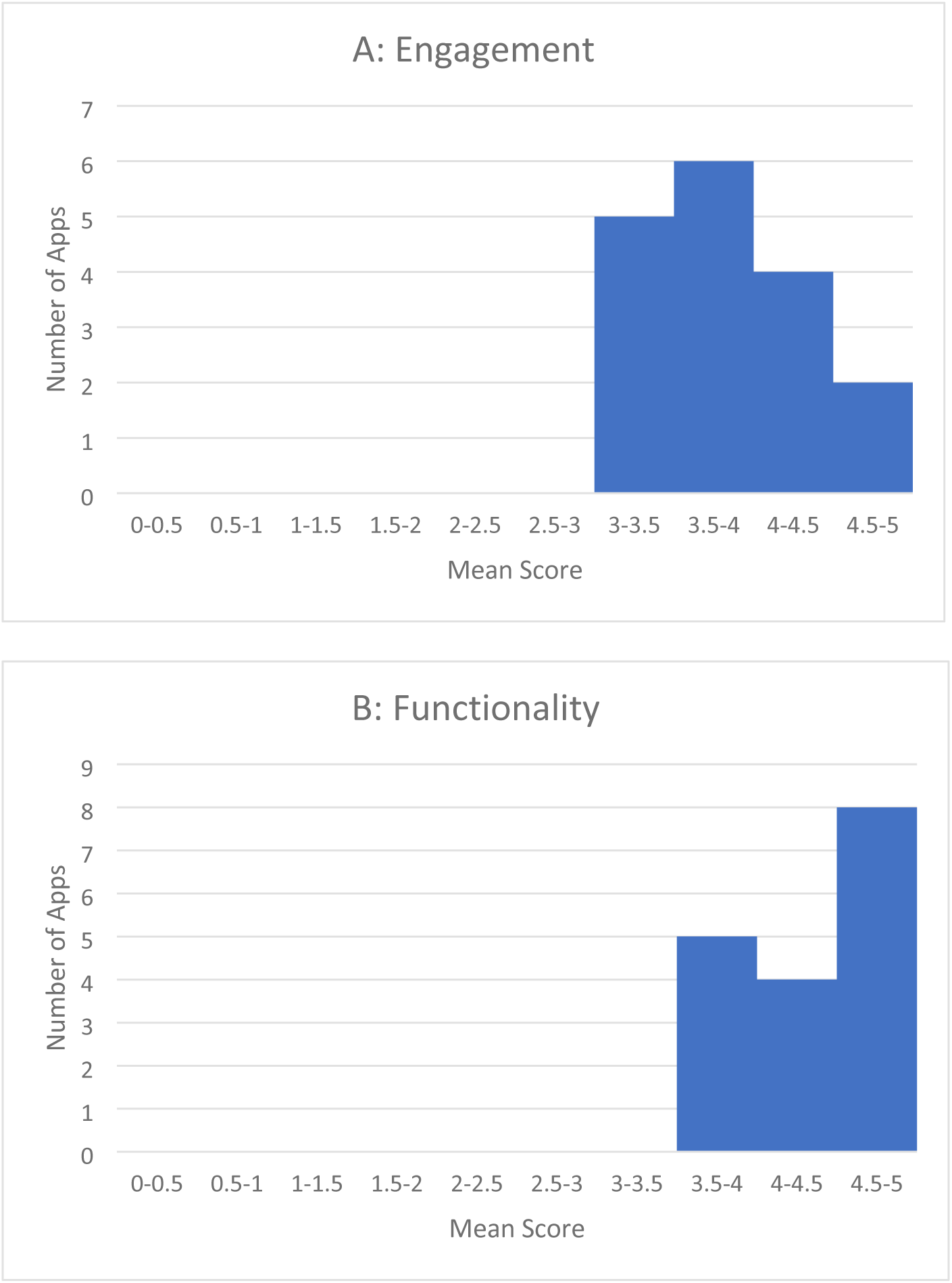
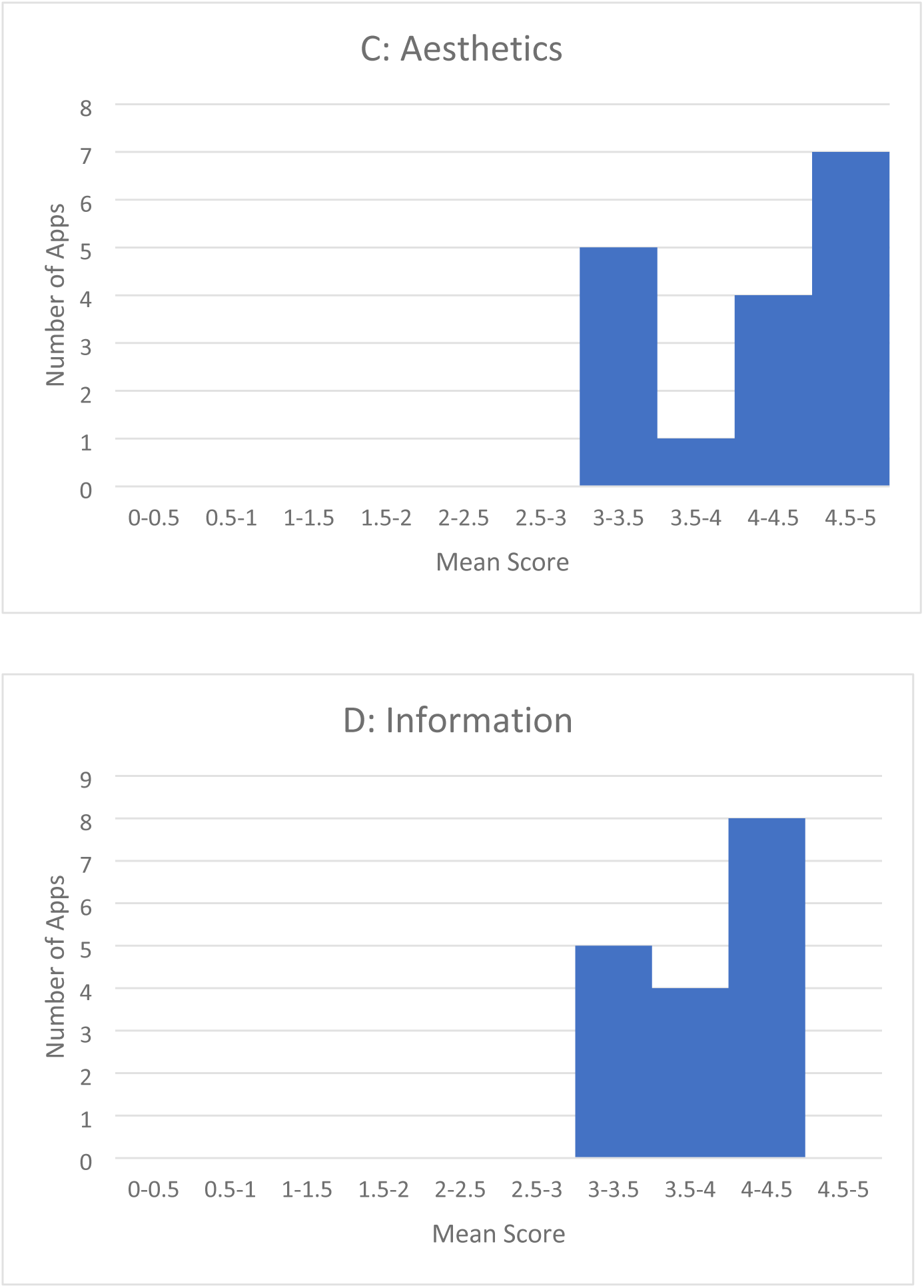
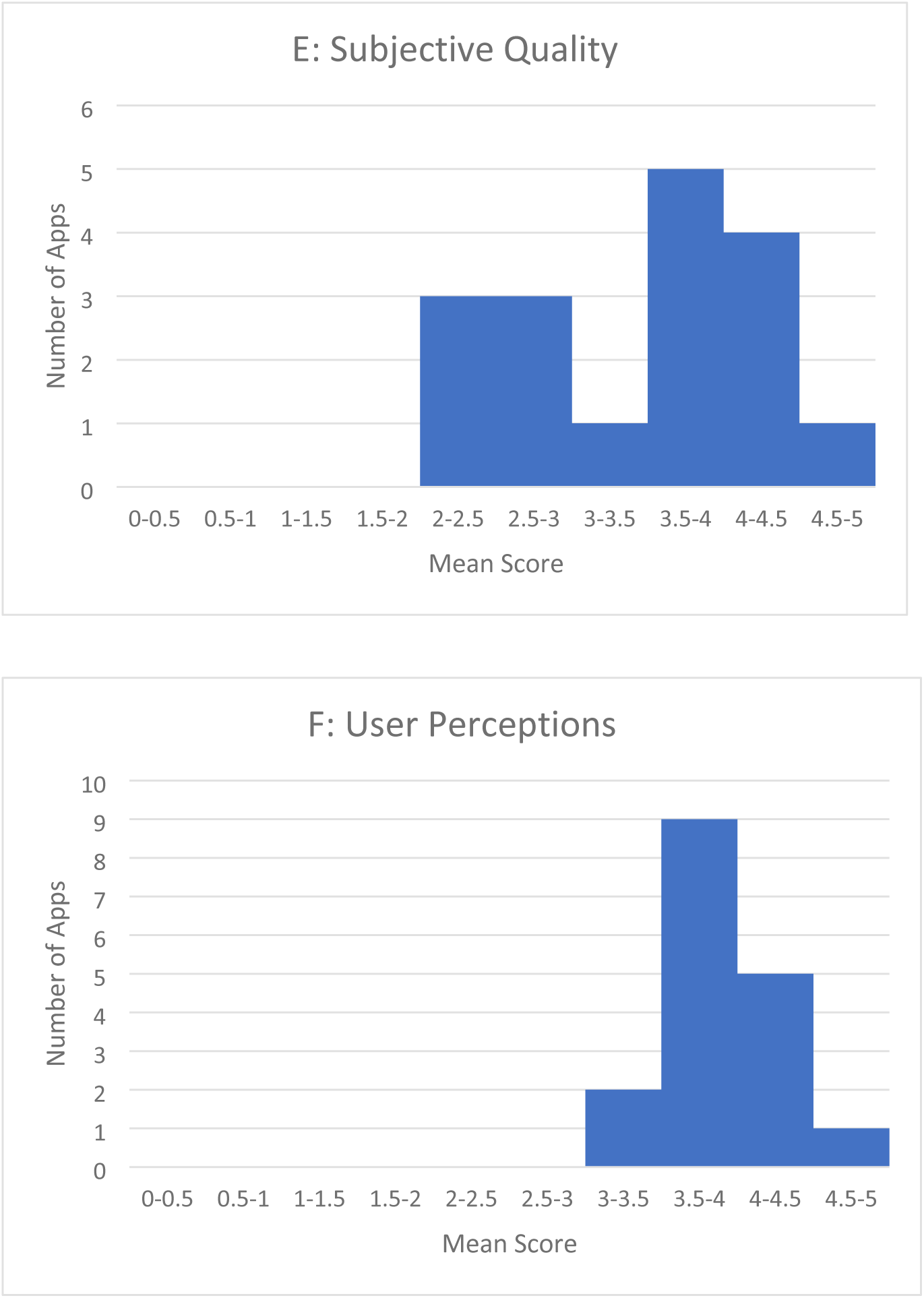
Mean MARS Ratings by Indicator.

### Exploratory linear regressions

An overview of exploratory linear regressions is provided in Table 5. For additional information, see Supplementary Table 3. The multiple linear regression of all MARS domains was the only analysis significantly correlated with app ratings (Adjusted R2 = 0.54, p=0.02). No other regressions showed significant correlations between ABACUS or MARS domains and app ratings.

**Table 5.**
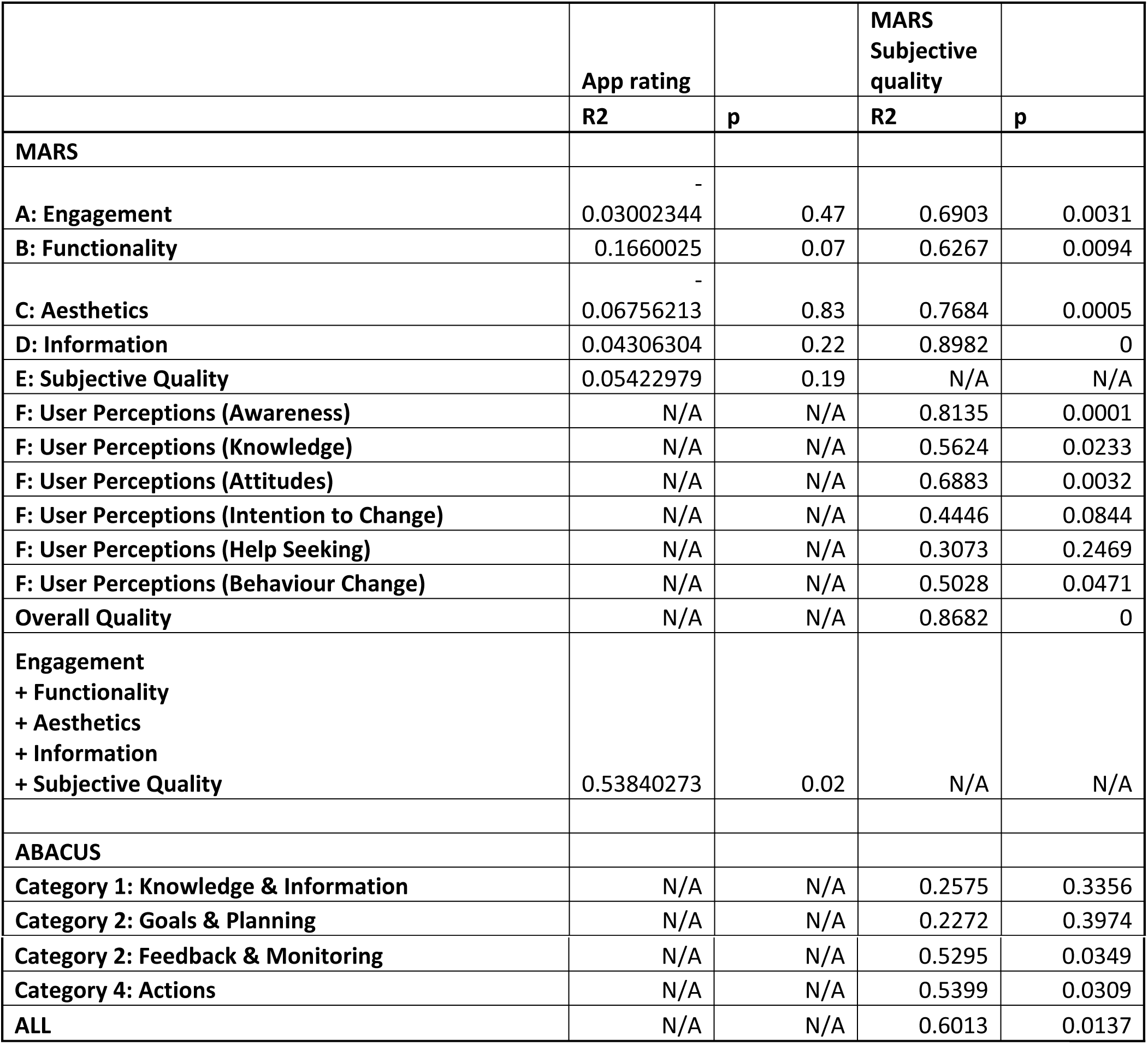
Summary of Linear Regressions.

With MARS subjective quality as the independent variable, mean scores for MARS engagement, functionality, aesthetics, information, as well as the overall quality score, showed significant positive correlations (R2 0.62-0.90, all P<0.05). App-specific user perceptions on awareness (R2=0.81, P<0.001), knowledge (R2=0.56, P=0.02), attitudes (R2=0.69, P=0.003), and behavior change (R2=0.50, P=0.047) also correlated significantly with MARS subjective quality. Lastly, ABACUS scores for feedback and monitoring (R2=0.53, P=0.03), actions (R2=0.54, P=0.03), and ABACUS total scores (R2=0.60, P=0.01) showed significant relationships with MARS subjective quality.

## Discussion

The purpose of this study was to explore behaviour change techniques, quality of features, and user ratings for interactive, publicly available apps promoting various PA modalities. An initial 600 apps were examined, though most were not interactive, required paid subscriptions or in-app payments, or required costly activity tracking hardware such as a Fitbit or Apple Watch and were thus excluded.

Accessibility of physical activities remains a barrier to many populations in particular, individuals living in lower socioeconomic status situations(37,38). Access to traditional PA, such as through gyms and recreational sports, may be cost-prohibitive or inaccessible for some populations(39) and thus non-traditional opportunities to improve PA may be instrumental for improving PA uptake. Yet, 73.2% of the global population have access to internet and 4.3 billion of the global population regularly use a smartphone(40,41), making digital apps an accessible solution to addressing low levels of PA.

As the number of internet users continues to rise globally, with 6.04 billion users(40), our sample focused on apps that were accessible to all individuals with internet access only. While none of the apps required user fees, many drove users towards paid features such as subscriptions, in-app purchases or wearables. All apps were commercially affiliated, with many apps promoting upgraded versions of the app via paid features or memberships. This demonstrates that freely available PA apps may only provide limited accessibility to PA uptake and adherence before users require monetary commitment.

A large proportion of apps were excluded due to lacking customization/interactivity (*n* = 284), not being PA-focused (*n* = 71), and requiring fees such as paid subscriptions (*n* = 47). The small number of apps meeting inclusion in the present study suggests that the current corpus of PA apps may be primarily intended to improve commercial gain, rather than or in addition to promoting PA for population health motives. Indeed, several of the included apps were associated with large, multi-national brands that may offer a PA app free of cost to users, as an extension of marketing efforts and brand recognition through interactive experiences and value-based activities such as education(42). For example, with over 10 million downloads, the Nike Training Club App has one of the lowest user rating scores in our sample. The Nike Training Club app included only 8 (38.1%) ABACUS items, explaining the comparatively low user ratings for the app. Regression analyses indicated that a greater number of ABACUS items was associated with improved subjective quality scores. Thus, an absence of ABACUS items related to facilitating PA behaviour change in the Nike Training Club app, coupled with implied commercial motives noted by the globally recognized Nike branding in all areas of the app, may have reduced user ratings. Prior research has shown that PA promotion with commercial motivations reduces user trust and perceived reliability(43) and has been associated with decreased perceptions of credibility and believability of health information(44). Users may have noted these commercial motives rather than focused support for goal-directed PA, resulting in reduced user ratings. This represents a may be that app developers are more focused on maximizing profit (i.e., product sales) compared with improving PA behaviour change notably, as any app designed for PA behaviour change would ideally intend for users to ultimately become sufficiently physically active over time, reducing the use of supportive PA technology.

All apps collected baseline information such as user age, gender, height and weight, presumably to allow the apps to provide tailored suggestions for PA based on exercise preferences commonly associated with these demographic criteria. These may include suggestions for reducing fat mass based on calculated body mass index, in cases where the app requested user height and weight. All apps also offered education about PA such as instructions on how to perform specific exercises by enabling users to see images, watch videos, or read instructions describing proper exercise technique. However, only two (11.7%) apps were consistent with PA guidelines (45) or demonstrated expertise in PA domains. As global PA trends suggest that 31% of the adult population do not meet current guidelines of 150 minutes of moderate-vigorous activity per week, (45) this gap may be a concern to PA and health professionals aiming to redress low-levels of PA.

User preferences for behaviour change techniques have shown that PA app users prefer to receive tips and instruction on PA(25). Research examining PA behaviour change suggests that individuals receiving reinforcing and positive feedback are more likely to engage in goal-directed behaviour, including those receiving remote or digital feedback(46,47). It is encouraging, then, that popular PA apps incorporate such behaviour change criteria, promoting PA uptake and in ways that are consistent with research exploring PA app user preferences(25). However, recent evidence suggests that improving PA may not be directive-driven rather, that individuals may benefit most from collaborative, practitioner/coach-facilitated goal setting and autonomy support for effective behaviour change(48).

While DeSmet et al(25) found that users ranked information about health consequences related to PA as a top preference in mobile apps, only one app (5.8%) in our sample included this feature. The target audience for many of the apps reviewed were young adults, where more proximal benefits of PA such as body image may be of importance(49); however, young adults may nonetheless be interested in PA to improve health(50). According to the ABACUS, this area addresses consequences related to continuing physical activities as well as consequences of discontinuing physical activities. Yet most apps focused on providing exercise instruction and advice to users. Those apps may have overlooked the relevance of sharing content related to behavioural outcomes, instead focusing on goal-directed outcomes. In other words, apps were generally centered around providing encouragement and feedback related to idealized goal achievement (e.g., body image ideals, improving strength). Yet many apps also included elements of behavioural consequences, such as PA consistency or daily step counts. While it is promising that apps allowed users to identify a variety of PA behaviours and outcomes of interest, it is important to consider whether achieving these outcomes is financially possible for all populations, given costs associated with tracking hardware such as smart watches to tally daily step counts.

Most (94.1%; n=16) apps included an option for goal setting, allowing users to identify their intended outcome expectations associated with app use. In some instances, goal setting was collected as baseline information, such that app users were required to identify at least one goal for app use (e.g., weight loss, increased muscle strength) as part of initial user set-up after downloading the app. Including goal setting as part of the user’s baseline information may also inform common app features, such as providing encouragement and offering feedback to the user: features that were identified in all included apps. Setting specific goals has been positively associated with physical activity among insufficiently active populations(51); however, for indirect PA outcomes such as self-efficacy and motivation, mixed and negative associations have been found with setting specific rather than general goals(51). It may be that impactful goal setting requires attention to an individual’s PA levels, as well as cognitive influences on PA, rather than the singular focus on physical outcomes found in most apps.

The ability for a user to self-monitor via an app has also been identified as an important feature for PA(25). Fourteen (82.4%) apps included a feature to allow users to track their activity either through self-report or direct measures, such as a smart watch. Specifically, trackable activities included recording daily steps/exercise, or connecting a wearable (optional) for behaviour tracking(24). Regression analysis also showed a positive relationship between *Feedback and Monitoring* with MARS *Subjective Quality* scales. Self-monitoring, including daily step counting, has demonstrated effectiveness for improving PA(52,53). Specifically, interventions that involve PA self-monitoring in combination with other behaviour change techniques, such as goal-setting or providing feedback, may improve PA uptake (52,53). While self-monitoring may be an effective approach to improving PA uptake, this does not contribute to long-term effects. Self-monitoring, including the use of wearable devices, does not appear to improve PA maintenance beyond three months (52). The ABACUS domain *Actions* also showed a moderate relationship with MARS *Subjective Quality*. The *Actions* domain focuses on habit formation, practice of daily activities, and restructuring physical and social environments to support behaviour change(24). Yet behaviour change techniques included in the ABACUS are focused on behaviour uptake rather than adherence. It may be that separate approaches are necessary for maintained PA, and diverse approaches to addressing PA uptake and adherence are needed. While the PA uptake focus of apps currently available to users addresses a relevant and timely issue, more attention is needed to develop supportive tools that help users achieve PA adherence over the long term.

The MARS allowed us to explore the PA apps for engagement, functionality, aesthetics, and information, as well as two separate scales for overall and subjective quality. Apps scored highest on functionality, suggesting that apps were usable, functional, navigable, intuitive, and easy to use.

Functionality and ease of use have been identified as key factors in user acceptability of digital health technology and may be a way users engage with digital health tools(54). Subjective quality, a score assigned by the research team to each app, scored lowest, and overall, apps were not recommended by the researchers for promotion and use for PA. To explore the relationship between MARS domains and app user ratings, linear regressions were computed; however, only aggregated MARS domains predicted app user ratings, explaining 54% of the variance in app user ratings. This finding suggests that individual MARS domains do not account for user perceptions in app rating scores. Other factors, such as subjective norms or reduced barriers, may also be influential. Users looking to engage with a PA app based upon recommendations from important others or due to ease of access (i.e., cost) may contribute to favorable user ratings, which may help to explain the generally favorable user ratings across all apps. This result may also reflect that apps often request user feedback ratings relatively soon after apps have been downloaded and users are in a ‘honeymoon phase’ of use with a new app, or idealized goal-directed behaviour. It may be that new features, information, and idealized outcome expectations may contribute to high user ratings for apps initially, prior to PA drop-out.

Despite behaviour change content analyses examining PA apps, this is the first study, to our knowledge, exploring PA apps that are both interactive and free of costs to users. As digital technology continues to evolve, app developers have begun to incorporate artificial intelligence (AI) in the conceptualization and design of mobile apps(55). Given the rapid increase and interest in PA apps that deliver interactive features such as feedback, monitoring, and personalization, the present study is also the first known study to exclusively examine PA apps incorporating at least one of these interactive features, which may include AI-driven functionality. Results from this study may support the development of PA apps that address known user preferences, while simultaneously adhering to evidence-based behaviour science recommendations. As such, this work strengthens our understanding of the overlap, or lack-thereof, between user preferences and behaviour science recommendations for PA apps. Use of the ABACUS and MARS frameworks as app-specific, validated tools is also a strength of this study, as validated codebooks were specific to behaviour change and app features. The availability of these validated tools improves the specificity of our findings and provides a behaviour-focused lens through which the apps were examined.

While there are several strengths to this research, there are also some limitations. The digital space is limited by temporality. Many PA apps fail to produce meaningful engagement with users and thus were neither returned in our initial searches nor captured in our data collection. This temporality may also limit replicability of this study, as PA apps may only have staying power when coupled with large, profitable brands or developers with broad user reach. Regarding data collection replicability, apps may have been ‘bumped’ to be returned as a top-ranking app by the search if app developers paid app stores for higher search return rankings. Both Google Play and Apple iTunes stores have similar yet limited user ratings ranges, which limited analyses and variance reported by user experiences. Similarly, the MARS Subjective Quality ratings may have biased other aspects of MARS ratings, as this scale is rated by research team members knowledgeable in behaviour science and may not align with public users’ considerations when rating apps. Results from this environmental scan were exploratory in nature and analyses in particular regression models reported here should be interpreted with caution. Finally, it may be the case that other popular PA apps exist but were not included in the present study if they were not English-based apps. Due to these limitations, the findings from this study are based on a small sample of PA apps which, at the time of data collection, were the only known zero-cost, interactive PA apps available on both Google Play and Apple iTunes systems.

## Conclusion

The ABACUS and MARS are app-specific scales which may identify key behavioural and quality features of apps for their potential to improve PA among adults. Findings from this study suggest that while all PA apps did incorporate at least some behaviour change techniques, few incorporated a majority of evidence-supported techniques. Nonetheless, all apps were free of cost to users, and publicly available, a key feature for improving equitable access to PA opportunities among all interested populations. While examining user ratings suggested that PA behaviour science and user preferences for PA apps are not yet aligned, user ratings were generally positive. Future work should examine how best to address PA using digital technology through approaches that are informed by behaviour change evidence, preferred by users, and can be used without cost, to equitably improve PA uptake and adherence and ultimately health outcomes associated with PA.

## Data Availability

All data are available within the publication. A data file is also deposited to the MRU Data Repository on Dataverse as Ori, Elaine, 2026, "Physical Activity Apps 2024".

https://doi.org/10.5683/SP3/LVSZ0I

